# Latent Therapeutic Demand of Immunoglobulin therapies in neuropathies – chronic inflammatory demyelinating polyneuropathy (CIDP), Guillain–Barré syndrome (GBS) and multifocal motor neuropathy (MMN) in the United States

**DOI:** 10.1101/2020.05.27.20114579

**Authors:** Megha Bansal, Albert Farrugia

**Affiliations:** 710 Medtronic Parkway, Minneapolis, MN, 55432-5604 USA; Faculty of Medicine and Health Sciences, University of Western Australia, 17 Monash Ave, Nedlands WA 6009, Australia

## Abstract

Chronic inflammatory demyelinating polyneuropathy (CIDP) is a neurological disorder characterized by progressive weakness and impaired sensory function in the legs and arms. Guillain-Barré syndrome is a disorder in which the body’s immune system attacks part of the peripheral nervous system. The first symptoms of this disorder include varying degrees of weakness or tingling sensations in the legs (NIH). Multifocal motor neuropathy is a progressive muscle disorder characterized by muscle weakness in the hands, with differences from one side of the body to the other in the specific muscles involved (NIH). We have modeled a latent therapeutic demand (LTD) of IVIg for CIDP and similar neuropathies in the US. We used the decision analysis methodology similar to the methods used by Stonebraker et al^1^ for modeling LTD of IVIg. The model is based on the relationships of the epidemiological and clinical factors. Most of the usage patterns and dosage level of albumin are according to the epidemiological studies and clinical trials. The model is built in Microsoft Excel. The analysis is conducted based on oneway sensitivity analysis and probabilistic sensitivity analysis. The demand in terms of grams per 1,000 inhabitants is calculated depending on the treatment schedule and the prevalence of the disease. The model for CIDP has eight variables including prevalence of CIDP, patients using IVIg, dosage and treatment patterns. The annual demand of IVIg is based on initial treatment of 24 weeks followed by a maintenance period, with lower dosage and frequency of treatment for another 24 weeks^2^. The model for GBS has eight variables with a loading dose for 3-6 days followed by a second dose in case of relapse. The model for MMN has nine variables. It has a loading dose followed by maintenance dose every 1-6 weeks depending on the clinical factors of the patient. On an average, IVIg use was calculated as 100 gms, 5.6 gms and 35 gms per 1,000 inhabitants for CIDP, GBS and MMN, respectively, in the US annually.

## Introduction

Chronic inflammatory demyelinating polyneuropathy (CIDP) is a neurological disorder characterized by progressive weakness and impaired sensory function in the legs and arms. Treatment for CIDP includes corticosteroids such as prednisone, which may be prescribed alone or in combination with immunosuppressant drugs. Plasmapheresis (plasma exchange) and intravenous immunoglobulin (IVIg) therapy are effective^3^. The US Food and Drug Administration (FDA) approved IVIg for use in CIDP in 2008^4^. We have modeled a latent therapeutic demand (LTD) of IVIg for CIDP in the US.

Guillain-Barré syndrome (GBS) is a disorder in which the body’s immune system attacks part of the peripheral nervous system. The first symptoms of this disorder include varying degrees of weakness or tingling sensations in the legs (NIH). The treatments include plasmpheresis and IVIg therapy^3^. IVIg benefit is uncertain in children but administration is considered by clinical experts. It is recommended for adults by AAN to lessen the immune attack on nervous system^3^. IVIg is approved by FDA for GBS^3^.

Multifocal motor neuropathy (MMN) is a progressive muscle disorder characterized by muscle weakness in the hands, with differences from one side of the body to the other in the specific muscles involved (NIH). MMN is a chronic disease and requires ongoing treatment of IVIg^3^.

## Methods

### Model Structure

We used the decision analysis methodology similar to the methods used by Stonebraker et al^1^ for modeling LTD of IVIg in neuropathies. The variables used in the model are shown in **Tables 1 and 2**. The model is based on the relationships of the epidemiological and clinical factors shown in the influence diagram **(Fig 1)**. Each disease follows a different treatment schedule and depending on the incidence of the disease the demand in terms of grams per 1,000 inhabitants is calculated.

**Table 1:**
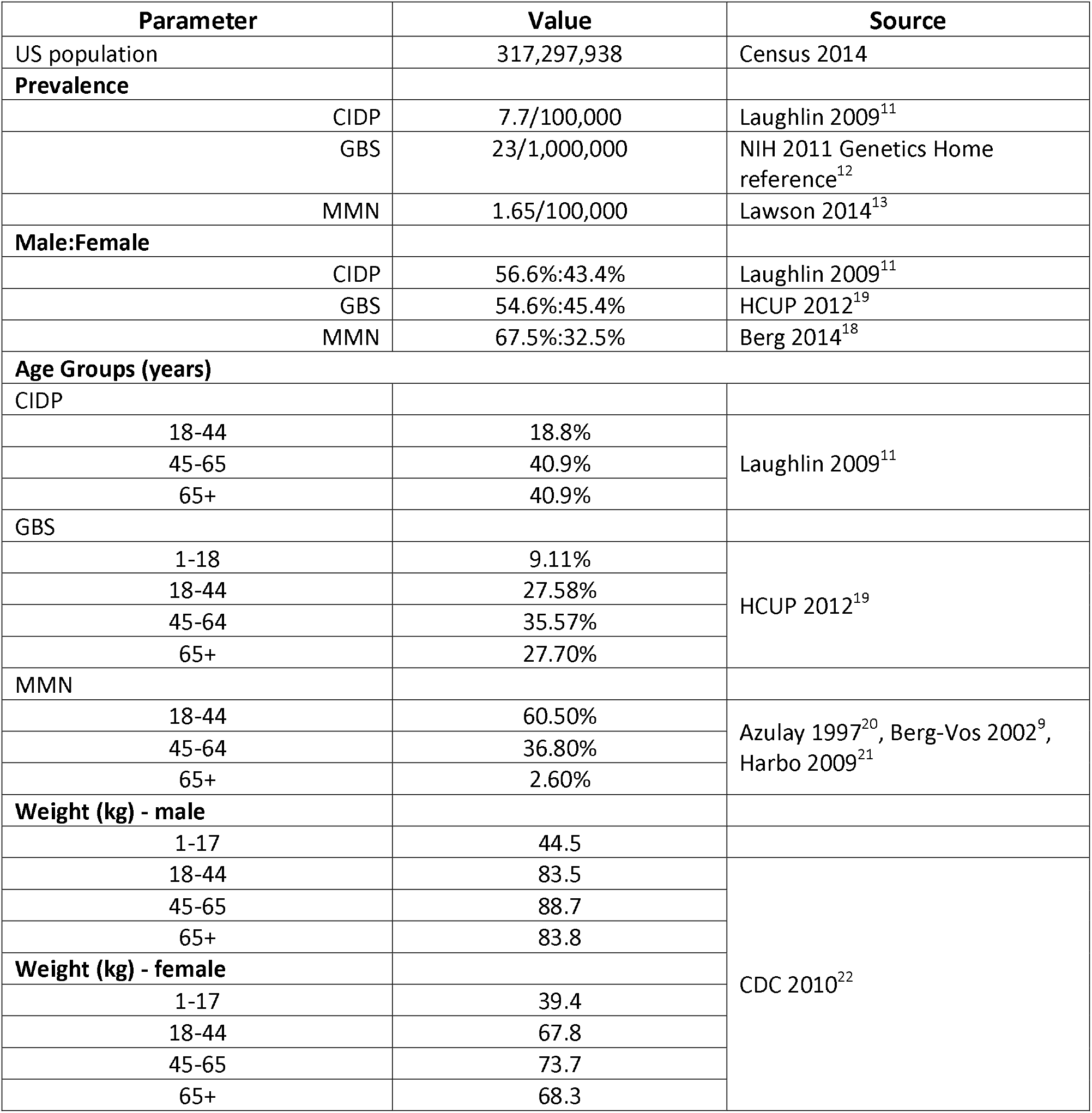
Population variables in the model

**Table 2:** Treatment patterns for CIDP, GBS and MMN

| Parameter | Value | Source |
| --- | --- | --- |
| **Treatment of CIDP** |  |  |
| Patients treated with IVIg in the initial period | 59% | Laughlin 2009 (table 1)<sup>11</sup> |
| Patients treated with IVIg in the extension period | 45.3% | Querol 2013<sup>15</sup> |
| Time period of IVIg administration | 24 weeks – initial period; 24 weeks – extension period | Hague 2008 ICE study<sup>2</sup> |
| Volume of IVIg used per person in the initial period | 2g/kg over 2-4 days and then 1 g/kg over 1-2 days | Hague 2008 ICE study<sup>2</sup> |
| Volume of IVIg used per person in the extension period | 1g/kg over 1-2 days | Hague 2008 ICE study<sup>2</sup> |
| Frequency of dosage in the initial period | Every 3 weeks | Hague 2008 ICE study<sup>2</sup> |
| Frequency of dosage in the initial period | Every 6-8 weeks or 8-12 weeks or >12 weeks | Querol 2013<sup>15</sup> |
| Patients on low frequency of dosage in the extension period |  |  |
| 6-8 weeks | 33.3% | Querol 2013<sup>15</sup> |
| 8-12 weeks | 35.9% |  |
| >12 weeks | 30.8% |  |
| **Treatment of GBS** |  |  |
| % treated with IVIG | 86.9% | Oczko-Walker 2010<sup>16</sup> |
| % treated with IVIG for second dose (patients who relapse) | 12% | va Doorn 2010<sup>7</sup>, NBA 2012 Australian guidelines<sup>8</sup> |
| IVIG administration days | 3.5 days | Patwa 2012 AAN guidelines<sup>5</sup> |
| volume of IVIG used per person | 1.2 g/kg | Patwa 2012 AAN guidelines<sup>5</sup>; NBA 2012 Australian guidelines<sup>8</sup> |
| frequency of dosage | may be repeated in case of relapse or no response | va Doorn 2010<sup>7</sup> |
| **Treatment of MMN** |  |  |
| % treated with IVIG | 80.0% | Jinka 2014<sup>10</sup>; 80% respond to the treatment of IVIG - could not find the exact usage data |
| % treated with IVIG maintenance | 100.0% | assumption |
| Interval between IVIG dose | every 3.5 weeks | Berg-vos 2002<sup>9</sup> |
| volume of loading IVIG | 2 g/kg | Jinka 2014<sup>10</sup> |
| volume of IVIG for maintenance | 1.1 g/kg | Berg-vos 2002<sup>9</sup>; Berg 2014<sup>18</sup>, NBA Australia 2012 guidelines<sup>8</sup> |

**Fig 1.**
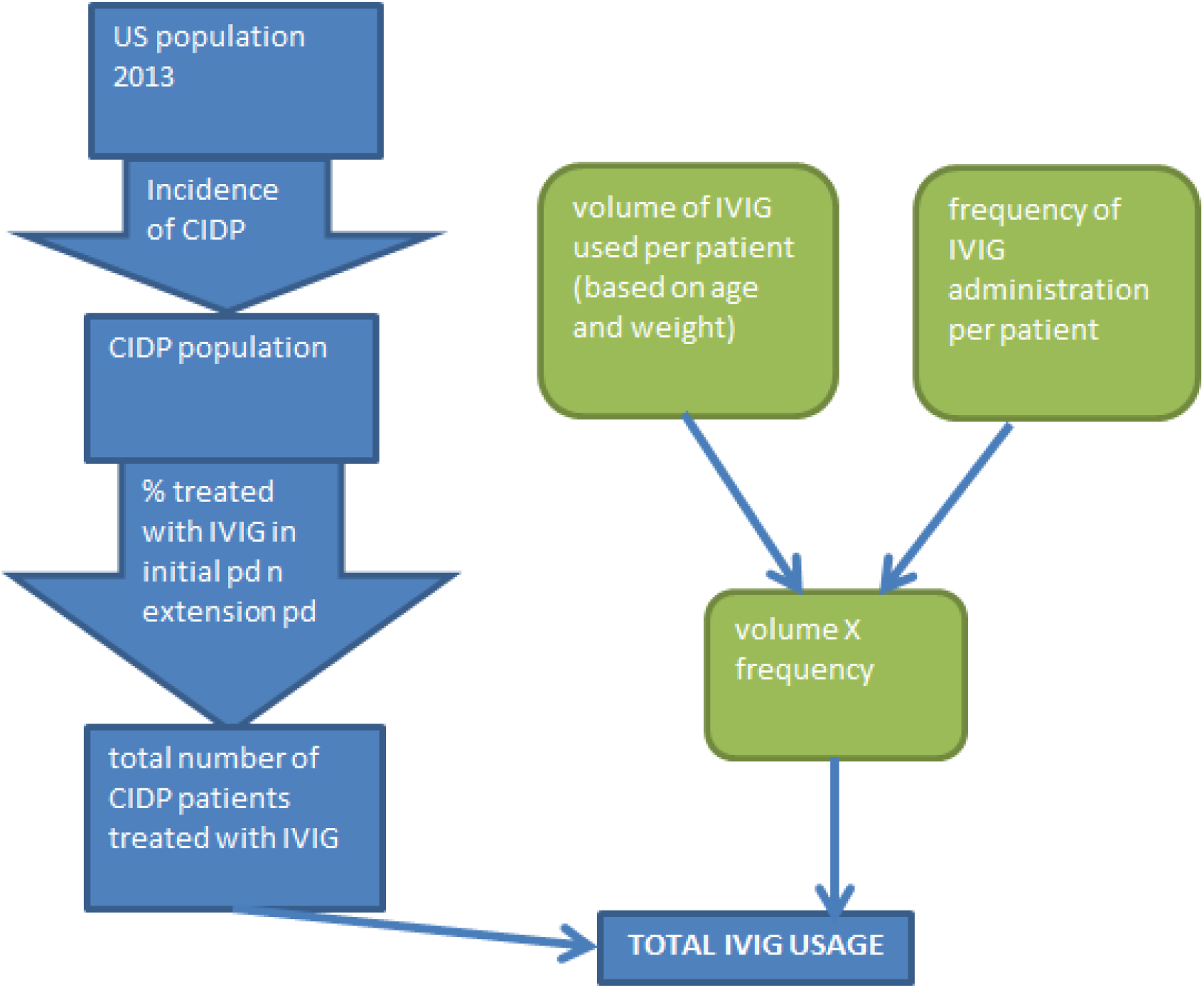
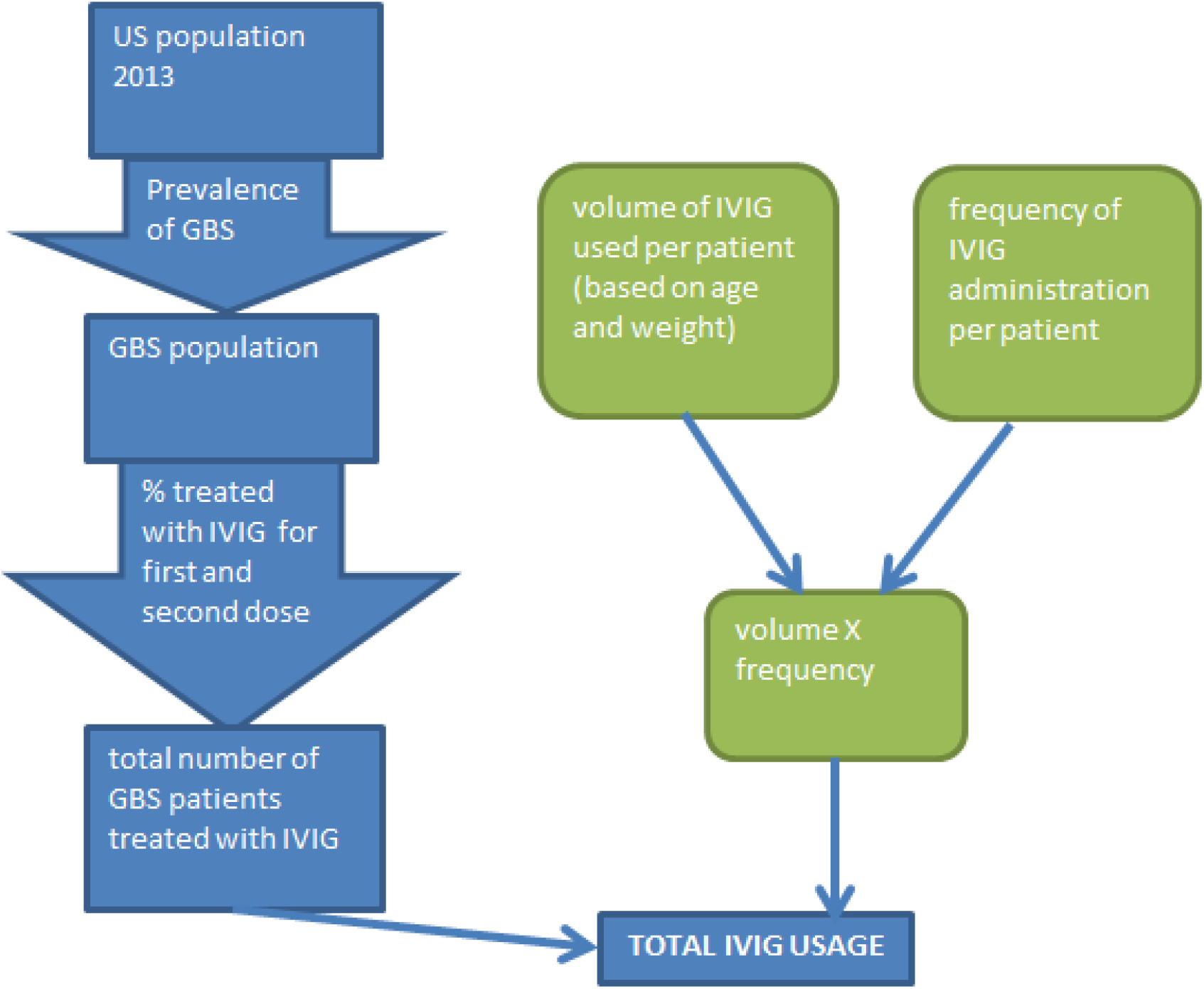
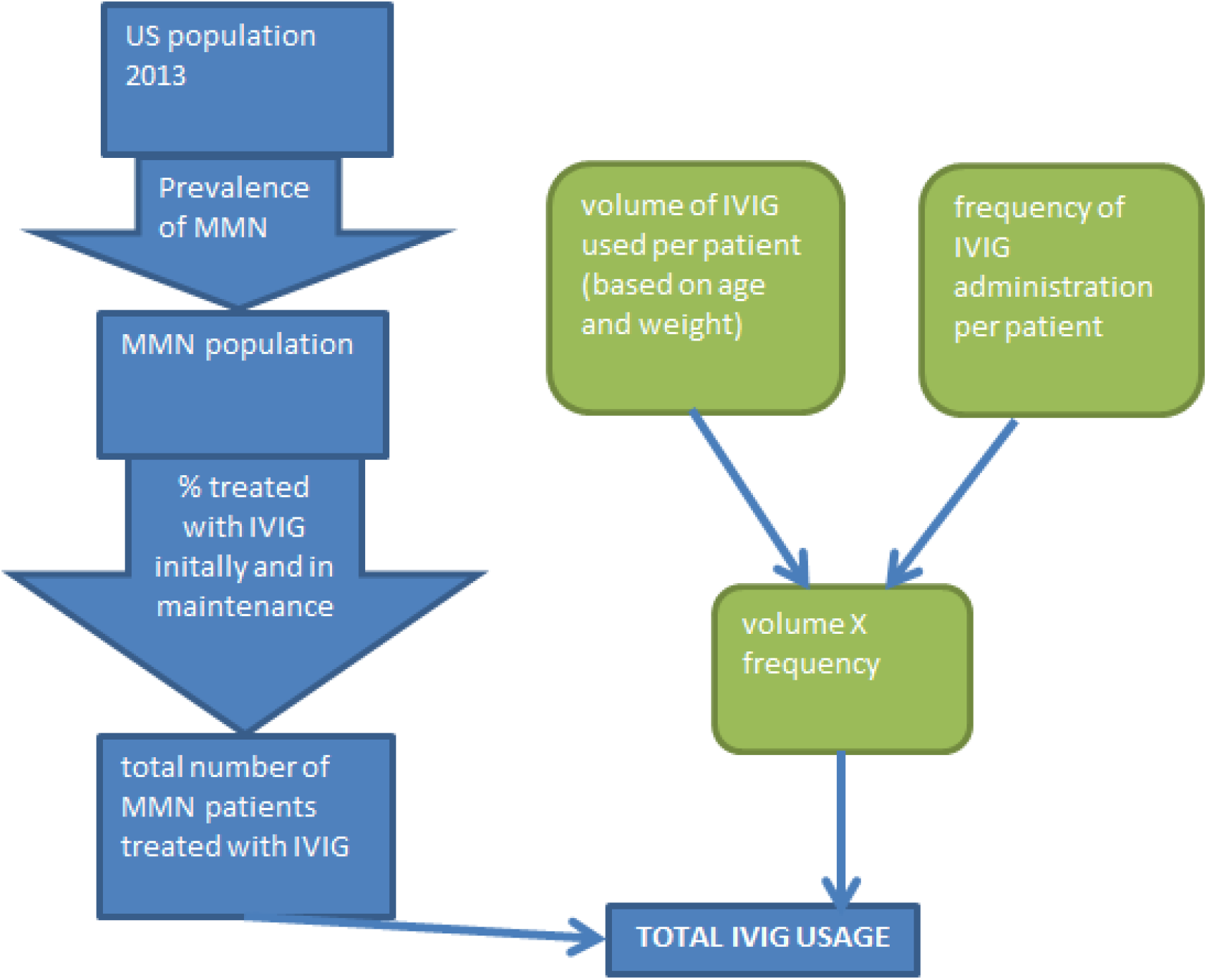
Influence diagram for use of IVIg in chronic inflammatory demyelinating polyneuropathy (CIDP) (a), Guillain-Barre syndrome (GBS) (b) and Multiple Motor neuron Neuropathy (MMN) (c)

The model for CIDP has eight variables including prevalence of CIDP, patients using IVIg, dosage and treatment patterns. The annual demand of IVIg is based on initial treatment of 24 weeks followed by a maintenance period, with lower dosage and frequency of treatment for another 24 weeks^2^. The American Association of Neurology (AAN) does not define any specific dosage levels for IVIg^5^. Thus, dosage was varied over a wide range to incorporate different levels provided by several sources^2,6^.

The model for GBS has eight variables with a loading dose for 3-6 days followed by a second dose in case of relapse^5,7,8^. The model for MMN has nine variables. It has a loading dose followed by maintenance dose every 1-6 weeks depending on the clinical factors of the patient^8–10^.

The model is built in Microsoft Excel. The analysis is conducted based on one-way sensitivity analysis and probabilistic sensitivity analysis. We have used probability distributions around variables as appropriate.

### Data Sources

#### Prevalence

A review of medical records in the US estimated the prevalence of CIDP as 8.9 per 100,000^11^. Other reports provided a varied range of 1.9 to 7.7 per 100,000 worldwide ^11^ (see Laughlin 2009 pg 40). Since the study done in the US showed much higher on a higher side compared to the worldwide estimation, we assumed 7.7 per 100,000 as the base case and conducted one-way sensitivity analysis around prevalence of 1.9-8.9 per 100,000.

According to National Institutes of Health, the prevalence of GBS is quite varied, 6 to 40 cases per 1 million people^12^. We have taken an average, 23 cases per 1 million as the base case. The one-way sensitivity analysis utilizes 6 and 40 cases per 1 million people as input values.

The prevalence of MMN is estimated from various sources to be 0.3-3 cases per 100,000 people^13^. The average of 1.65 cases per 100,000 people is used as the base case.

#### Patients treated with IVIg

CIDP is a rare disease and the review of medical records in the US identified 23 cases treated for CIDP in the Rochester Epidemiology Project^n^. Among these cases, 59% were treated with IVIg. Another retrospective study done in Massachusetts General Hospital identified 28.9% of patients being treated with IVIg in CIDP. We used a uniform distribution to vary the patients treated in the initial period.

In the maintenance or extension period, the CIDP patients that responded well to IVIg were infused with lower dosage of IVIg and at lower frequency to avoid a relapse. Multiple studies and trials showed the drug was well responded in 50-70% patients^3,14^. A retrospective study done in Spanish hospitals showed 45.3% of the patients responded well to the treatment and were assigned to the group with low frequency of infusions. This group was further divided among patients receiving drug at 6-8 week interval, 8-12 week and >12 weeks^15^.

A retrospective chart review study shows 86.9% of the patients receive IVIg either alone or in combination with plasmapheresis^16^. Out of these, patients who relapse are provided with a second dose of IVIg. The evidence shows 8% to 16% of the patients relapse^7,8^.

A review study^10^ for treatment of MMN reports 80% of the patients respond to IVIg. We used this figure (80%) as the number of people treated with IVIg in the US due to lack of exact data. Since treatment of MMN is an ongoing treatment, we assumed 100% of these patients are treated with IVIg.

#### Dose of IVIg

We noted the administration of IVIg for one year with 24 weeks on the initial period and 24 weeks in the maintenance period. Most of the trials and studies mentioned a loading dose of 2g/kg of IVIg in the initial period over 2-4 days. For the remaining period, a range of 0.4-2g/kg of IVIg is given every 3 weeks^2,6,17^. The AAN recommends IVIg and suggests the dosage, frequency and duration of treatment based on clinical assessment^5^. Since there were no criteria defined for dosage, we varied the dosage using a uniform distribution.

In the extension period, the same range of 0.4-2g/kg of IVIg was used but the frequency of infusion was reduced from 3 weeks to 6-24 weeks. The patients were divided in the groups of 6-8 weeks, 8-12 weeks, 12-24 weeks^15^.

A randomized controlled trial^5^ used 0.4g/kg of dosage for GBS over 3-6 days. The Australian guidelines recommended dosage to be 2g/kg over 2-5 days^8^. We used an average of 1.2g/kg over 4 days as the base case and varied the dose from 0.4 to 2g/kg over 3-6 days in one-way sensitivity analysis.

The evidence shows an induction dose of 2g/kg of IVIg is administered in MMN ^10^. The maintenance dose varies between 0.1-2g/kg every 1-6 weeks annually^8,9,18^.

#### Age, Gender and Weight of patients

In the cases where dose is administered per kg of the weight of patients, it is necessary to distinguish age and gender groups. The data for gender and age are obtained from the retrospective study conducted in the US ^11^. The age groups are classified as 18-44, 45-64, 65+ years of age. There was only one case of a 4 year old in the retrospective study^11^ for CIDP which we excluded so that the population is aligned with the adult population in most of the clinical trials^2^.

GBS inflicts both adults and children so we included another age group of 1-18 years of age to GBS. The proportions of patients in each age group were gathered from HCUP 2012 ^19^. Several follow up studies were used to populate the proportions in each age group in MMN^20,9,21^

The weight is obtained from a 2012 report by CDC^22^. They outline weight by age and gender. We used extended Swanson-Megill (ESM) approximation to obtain 10^th^, 50^th^ and 90^th^ percentiles and used it in probabilistic sensitivity analysis.

## Results

On an average, IVIg use was calculated as 100 gms, 5.6 gms and 35 gms per 1,000 inhabitants for CIDP, GBS and MMN, respectively, in the US annually. The one-way sensitivity analysis **(Table 3)** derived from the model showed the dose in the initial period and prevalence impacted the demand of IVIg the most **(Fig 2)**. The treatment frequency in the maintenance period affected the demand the least. The probability distributions of the LTD among all these neuropathies show a right skewed distribution **(Fig 3)**.

**Table 3:**
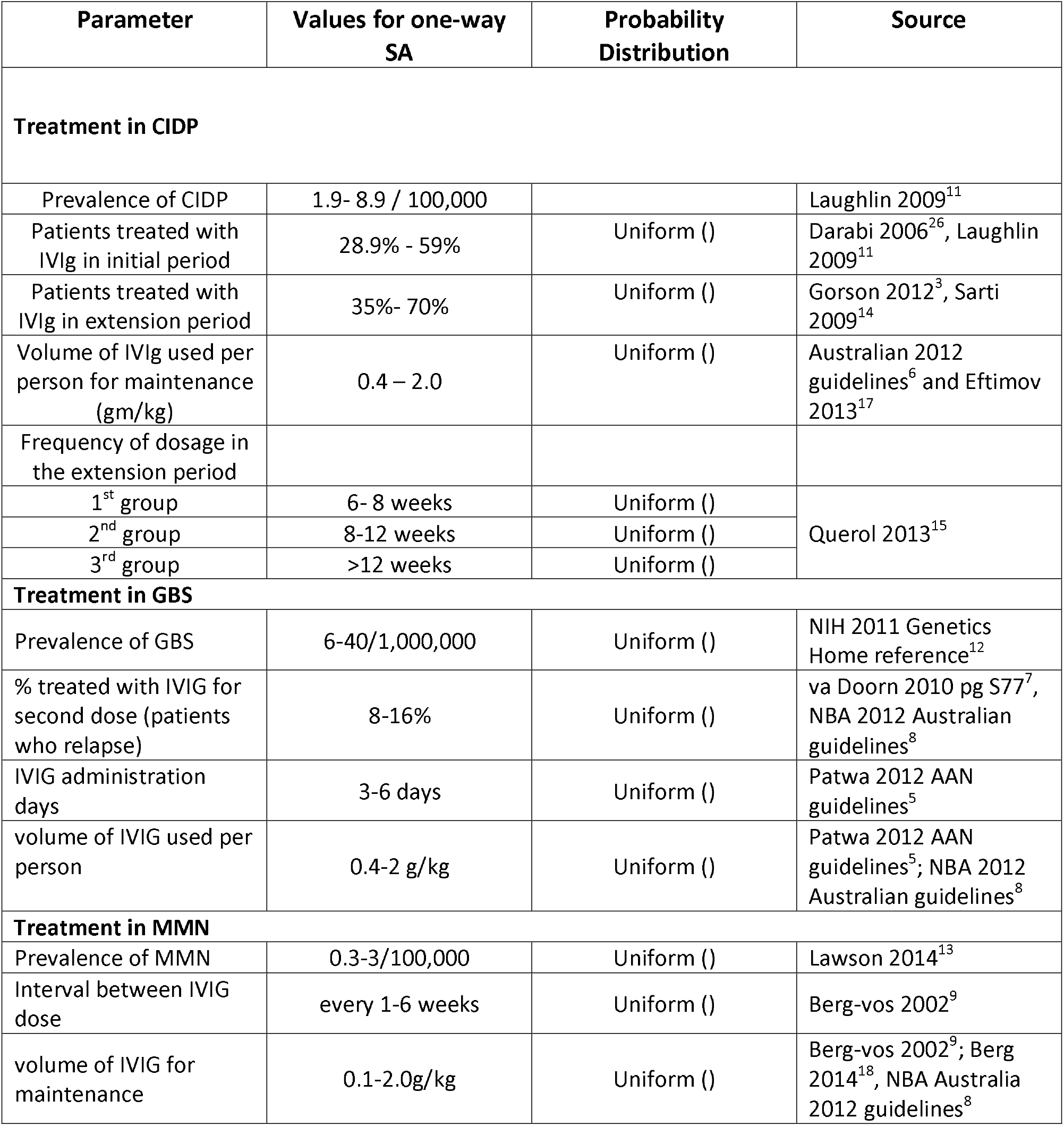
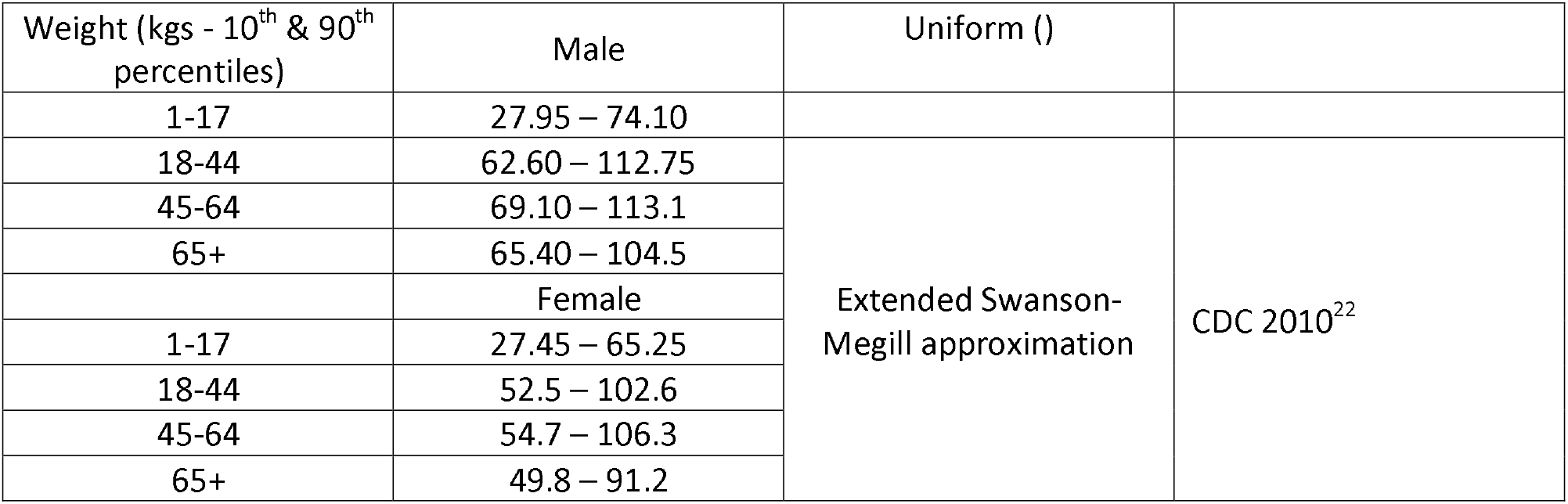
Variables used in the sensitivity analysis

**Fig 2.**
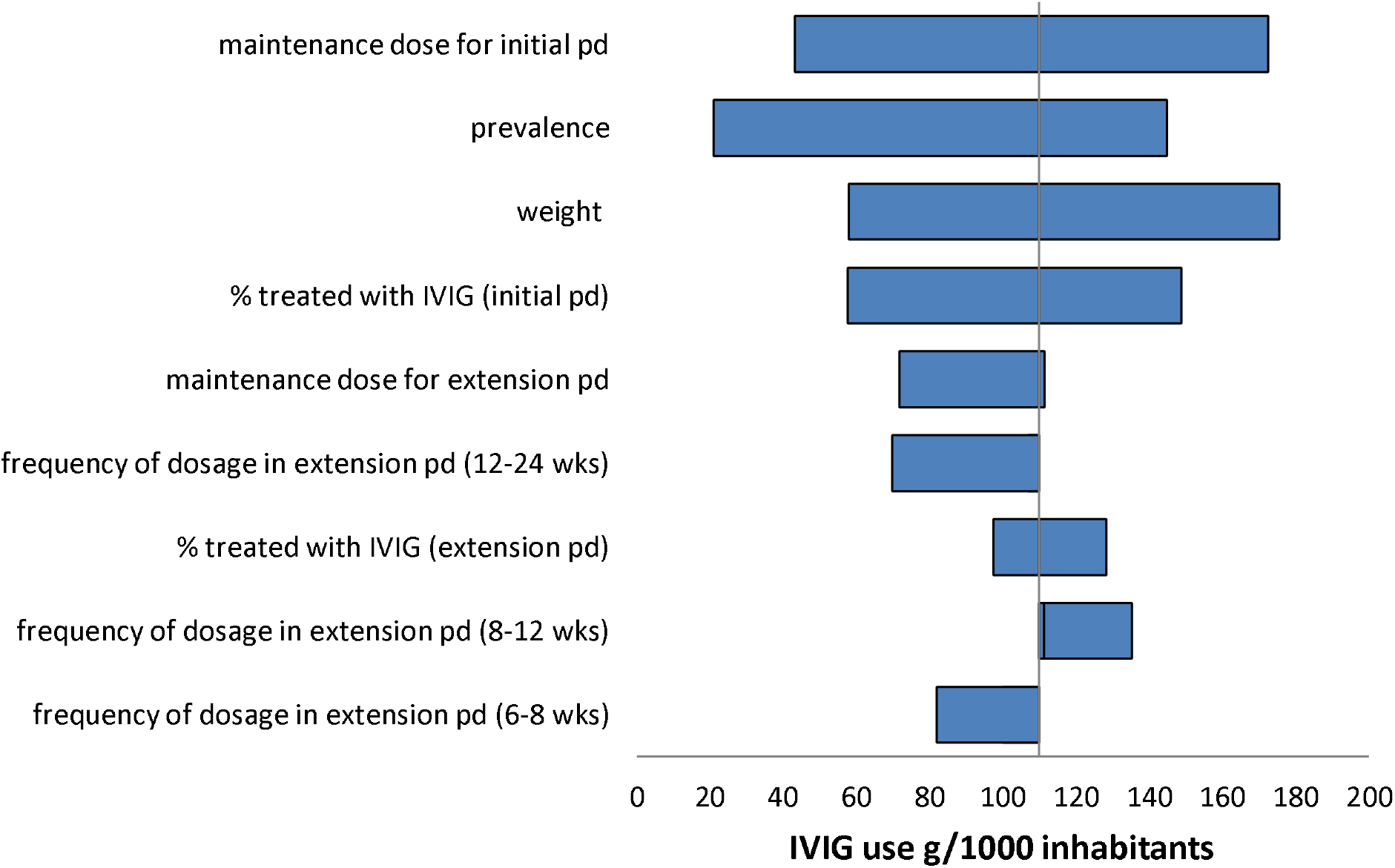
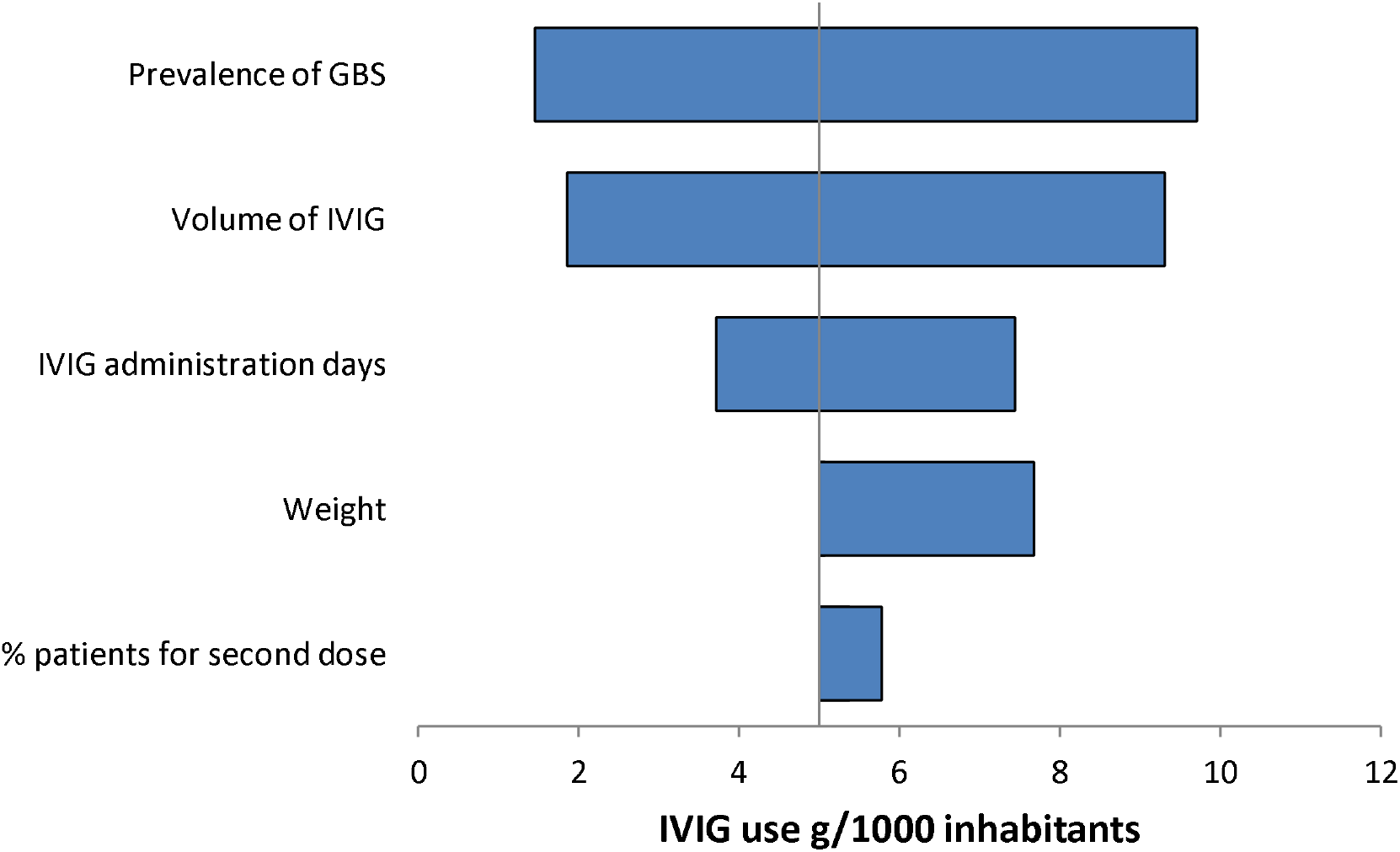
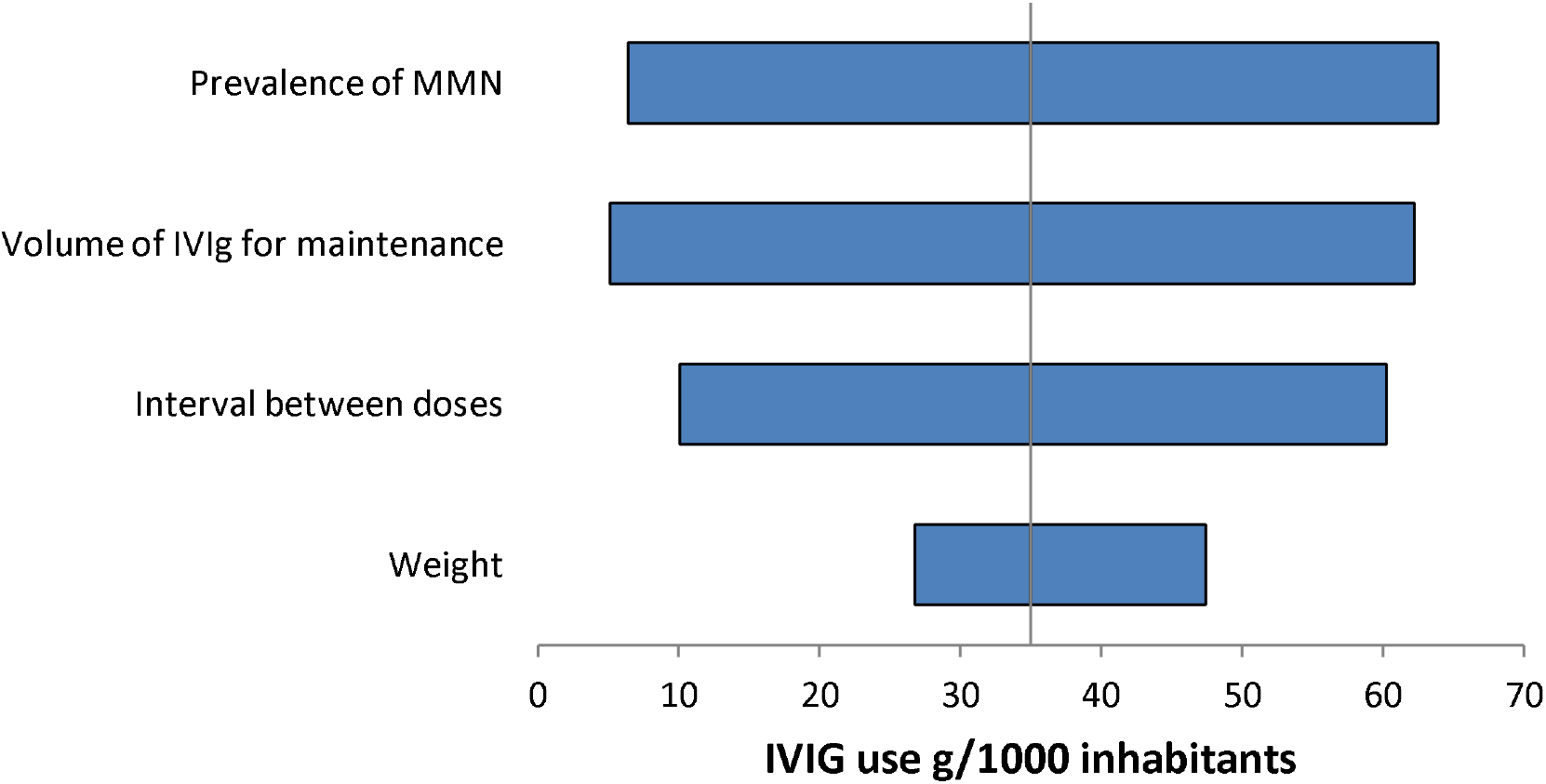
One-way sensitivity analysis (tornado diagram) - for variables included in the decision analysis model for CIDP (a), GBS (b) and MMN (c)

**Fig 3.**
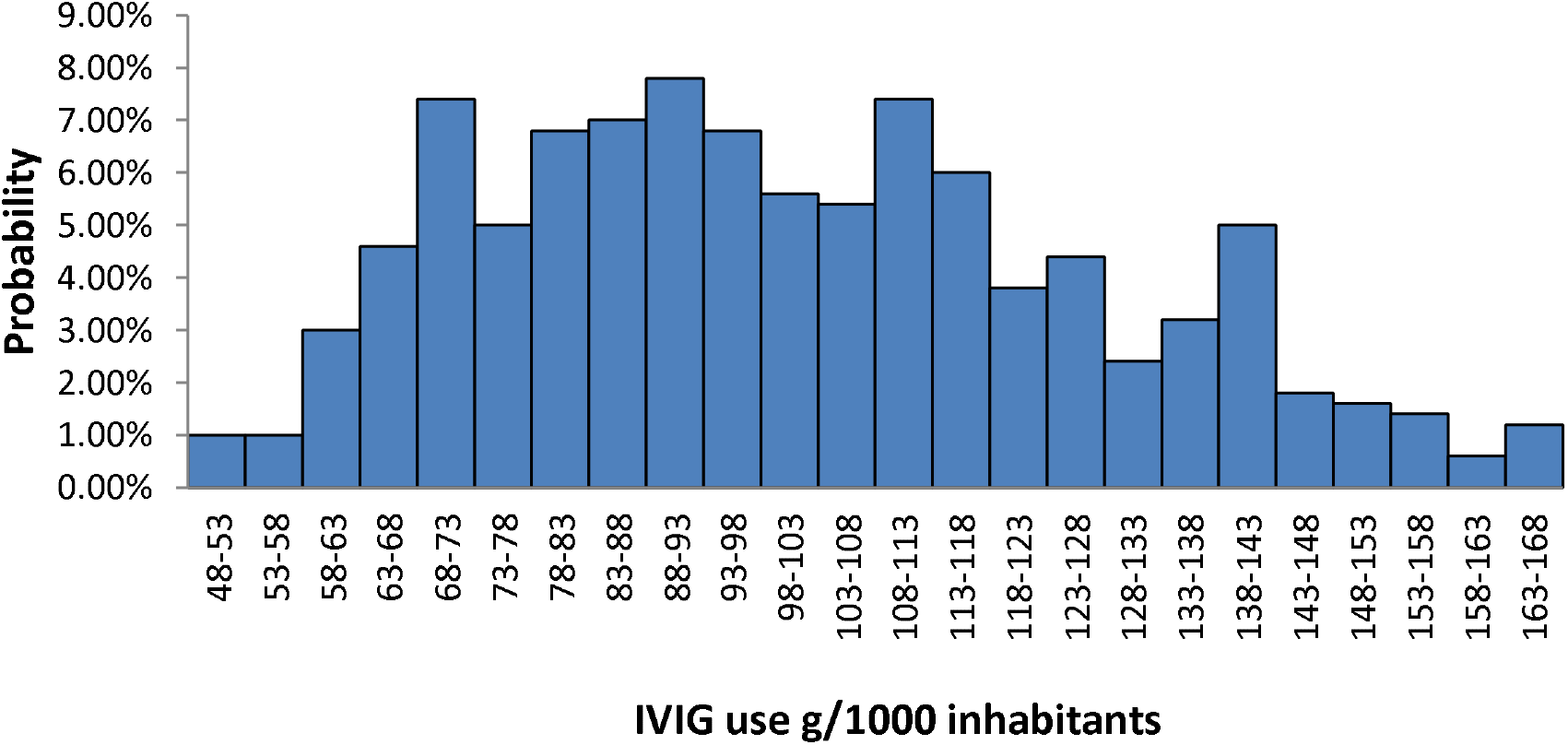
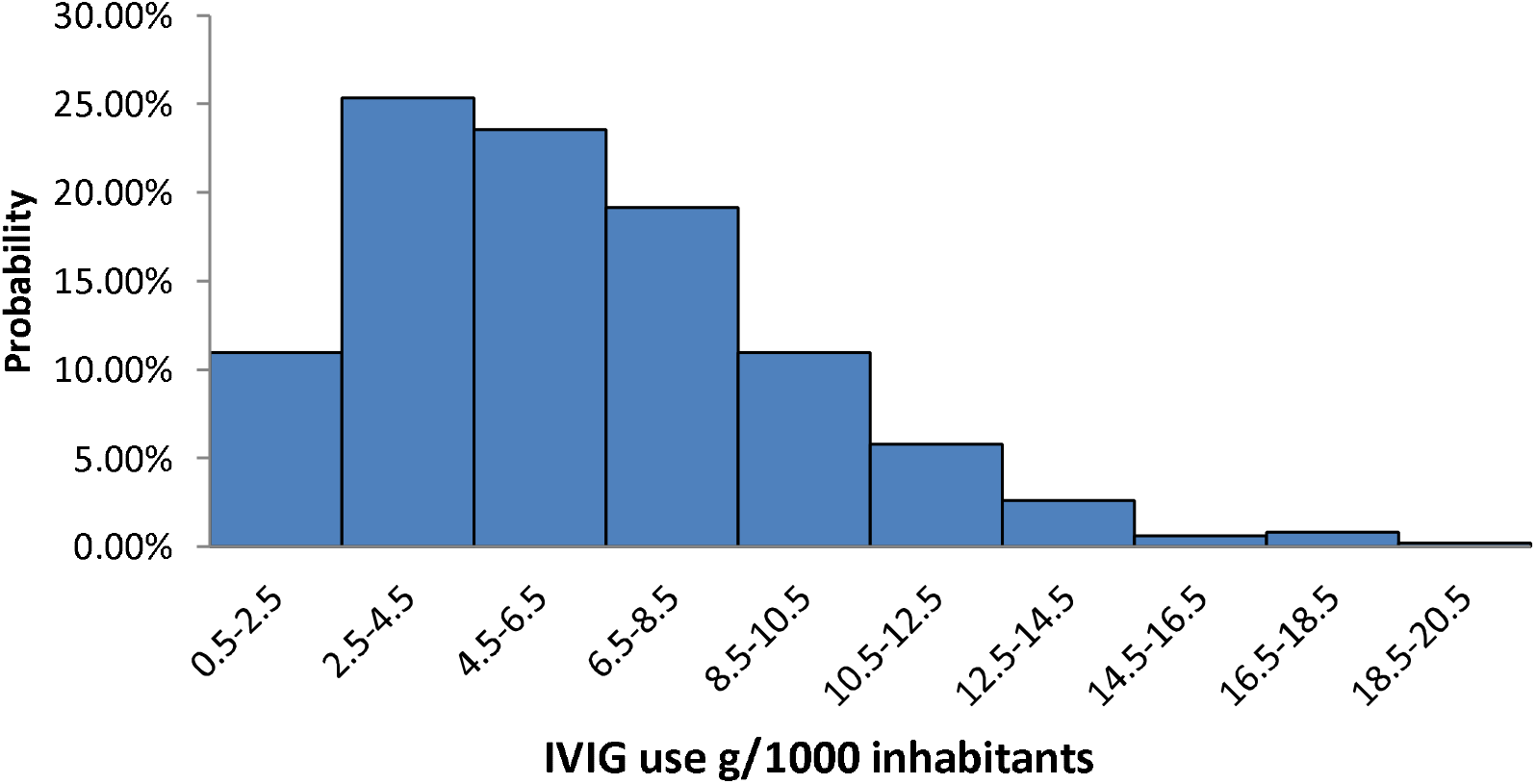
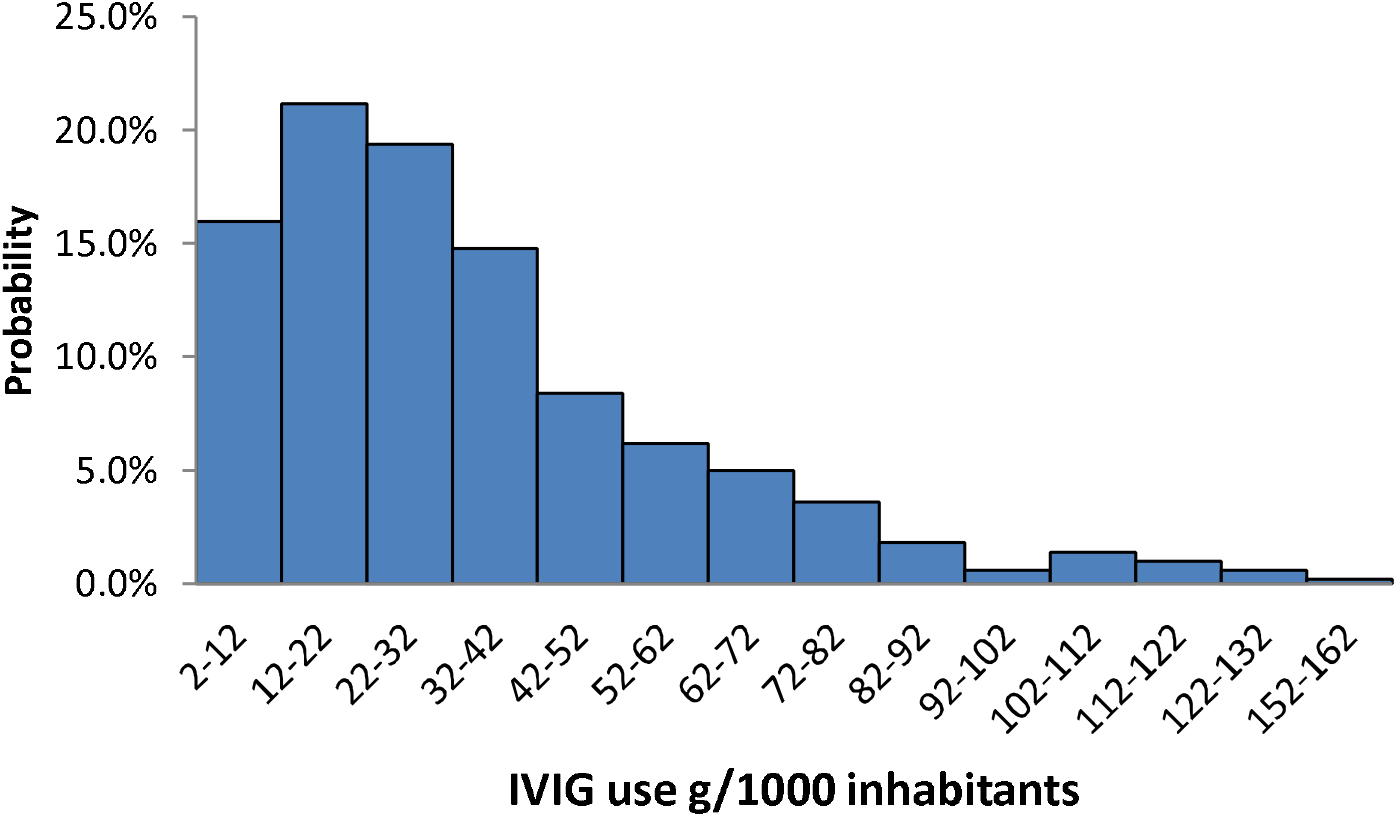
Distribution of IVIG use in the US, extracted from the model and estimated for patients with CIDP(a), GBS(b) and MMN (c)

## Discussion

The average demand and standard deviation for CIDP, GBS and MMN are 97.9 ± 24.5, 6.1 ± 3.2 and 34.4 ± 25.5 g/1000 inhabitants, respectively. The combined evidence based underlying demand for IG in these necrologies approaches, therefore, 140 g/1000 inhabitants. Combined with the estimated demand of 105 g/1000 population for the primary immune deficiencies^23^, this suggests that the amount of IG used currently in the countries using high amounts IG reflects the consumption expected from decision analysis modelling. As diagnosis and treatment access improve in other, lower consuming countries, the actual usage is expected to approach the levels estimated in these models. The global supply of IG is already under substantial stress^24^, and increased effort is required to collect the volumes of plasma needed to generate these amounts of IG. These pressures should heighten the search for alternatives to IG therapy, with some therapies already showing promise as candidates for treating the autoimmune neurological diseases assessed in the present study^25^. In the interim, we propose that decision analyses for estimating latent demand for therapies should be considered as useful tools in assisting policy makers and funders in planning access to medical interventions.

## Data Availability

All the data and references to this paper are freely available.

## Notes

### Competing Interest Statement

The authors have declared no competing interest.

### Funding Statement

No funding was provided for this article.

### Author Declarations

No research in human subjects was performed in this study

